# Building and sustaining trust across communities: Lessons from a large-scale, community-based cancer needs assessment in New York City

**DOI:** 10.1101/2025.11.20.25340674

**Authors:** A Rakhra, Y Yusuf, D Min, V Foster, S Sifuentes, A Kazmi, SC Kwon

**Affiliations:** Department of Population Health, NYU Grossman School of Medicine; Vilcek Institute of Biomedical Research, New York University Grossman School of Medicine

**Keywords:** community health, community engaged strategies, needs assessments, community-engaged research, community health workers

## Abstract

**Background:** Trust is central to healthcare engagement yet remains underexamined in the context of large-scale community needs assessments. The Cancer Community Health Resources and Needs Assessment (Cancer CHRNA) was developed and implemented to identify multilevel determinants for cancer prevention and disparities; and examine the structural and system levels factors influencing healthcare access and prevention behaviors across populations represented in New York City. This study examines how trust emerged as a dominant theme and the relational and technical strategies community health workers (CHWs) and community-based organizations (CBOs) used to establish and sustain trust during survey implementation.

**Methods:** Cancer CHRNA was implemented in community– and clinic-settings in nine languages: English, Arabic, Bangla, Chinese-Simplified/Traditional, Haitian-Creole, Korean, Spanish, Russian, and Urdu by bicultural and bilingual CHWs, in partnership with a network of CBOs. This qualitative process evaluation draws on data from CHW interviews, field notes, and a research team focus group. Analysis was guided by the Consolidated Framework for Implementation Research (CFIR), with secondary coding informed by Metz et al.’s theoretical model for trusting relationships, which distinguishes relational and technical strategies of trust-building.

**Results:** Three overarching themes related to trust emerged: 1) CHWs as trusted messengers embodying trustworthiness; 2) the role of CBO partnerships in enhancing trust; and 3) the development of sustained trust with both CBOs and community members. Across these themes, CHWs and CBOs employed relational strategies (authenticity, empathy, bi-directional communication, vulnerability) and technical strategies (cultural and linguistic concordance, demonstration of expertise, frequent interactions, responsiveness). These strategies activated trust, which in turn enabled successful recruitment and sustained engagement.

**Conclusion:** To our knowledge, Cancer CHRNA is the first needs assessment to assess cancer behavioral and social priorities in nine languages, providing a unique exploration of the role of trust within such an assessment. Findings demonstrate that culturally and linguistically concordant CHWs, working in partnership with trusted CBOs, were central to fostering trust across the relational and technical strategies of trust building and facilitating broad community participation. By highlighting trust as the mechanism underpinning recruitment success, this study offers practical insights for designing future multilingual, community-based assessments.

## Introduction

Trust is essential to delivering health care and improving health outcomes(1–3). Lack of trust in the healthcare system is a growing and complex problem with historical and contemporary roots involving individual and community encounters with the government, medical institutions, and health care providers. Trust can impact one’s utilization of medical care, medication adherence, continuity of care, and self-reported health status(3–7). As such, it is increasingly being recognized as a critical aspect of medical care and greater attention has been placed on understanding and improving it.

Community health workers (CHWs) are widely acknowledged as trusted messengers within their communities (8–12). The American Public Health Association (APHA) defines a CHWs as “frontline public health worker who is a trusted member of and/ or has an unusually close understanding of the community served, enabling them to act as liaisons between health and social services and the community.”(10) Their cultural and linguistic concordance, personal lived experiences, and deep community ties allow them to serve as bridges between healthcare systems and historically underserved populations (8–12).

Similarly, community-based organizations (CBOs) serve as liaisons for a community and provide educational, social, or related services to members of the community(8). Being embedded within communities, CBOs identify community priorities and understand the culture, language, community norms, and best strategies for outreach and engagement. For marginalized communities who report a lack of trust in medical institutions, CBOs can serve as a trusted source of health messaging and foster trust between individuals and healthcare systems (8). CBOs and community-engaged processes are key to building and sustaining trust in research collaborations(13). Despite their central role in community-engaged research, the specific processes through which CHWs and CBOs build and sustain trust during large-scale needs assessments remain underexamined.

Immigrant populations are particularly underrepresented in research and face well-documented disparities in cancer prevention and screening due to language barriers, limited access to care, and a lack of culturally tailored services (14–16). In New York City, where over one-third of residents are foreign-born (17), these challenges underscore the need for research approaches that are community-centered, linguistically accessible, and explicitly designed to build trust.

The Cancer Community Health Resources and Needs Assessment (Cancer CHRNA) was developed to address these disparities using a community-based participatory approach (CBPR). Bilingual and bicultural CHWs partnered with CBOs to co-develop the survey, identify recruitment sites, and lead multilingual data collection across nine languages. While the assessment aimed to identify determinants of cancer prevention, assess structural factors that influence healthcare, and document community priorities, our qualitative process evaluation revealed that trust emerged as a central theme across implementation.

To evaluate these dynamics, we applied the Consolidated Framework for Implementation Research (CFIR) (18) to examine multilevel contextual influences on implementation, and Metz et al.’s the theoretical model for trusting relationships and implementation (19), to analyze relational and technical strategies CHWs and CBOs used to build and sustain trust.

This paper aims to: 1) describe how trust emerged across qualitative data sources; 2) highlight the relational and technical strategies used by CHWs and study partners to build and sustain trust; and 3) discuss how these findings can inform the design and implementation of future community-based needs assessments seeking to engage populations with lower levels of institutional trust.

## Methods

### Needs Assessment Overview

The Cancer CHRNA was implemented from July 2021 to November 2022 through a collaborative effort involving multiple departments within a major metropolitan New York-based academic healthcare system, including a nationally recognized cancer center and one of the largest Federally Qualified Health Centers (FQHCs) in the U.S. Cancer CHRNA was implemented in clinic and community-settings in nine written languages: English, Arabic, Bangla, Chinese-Simplified and Traditional, Haitian-Creole, Korean, Russian, Spanish, and Urdu (see Table 1). To our knowledge, the Cancer CHRNA is the only needs assessment that assessed cancer-related social and behavioral priorities and risk factors in nine languages.

**Table 1.**
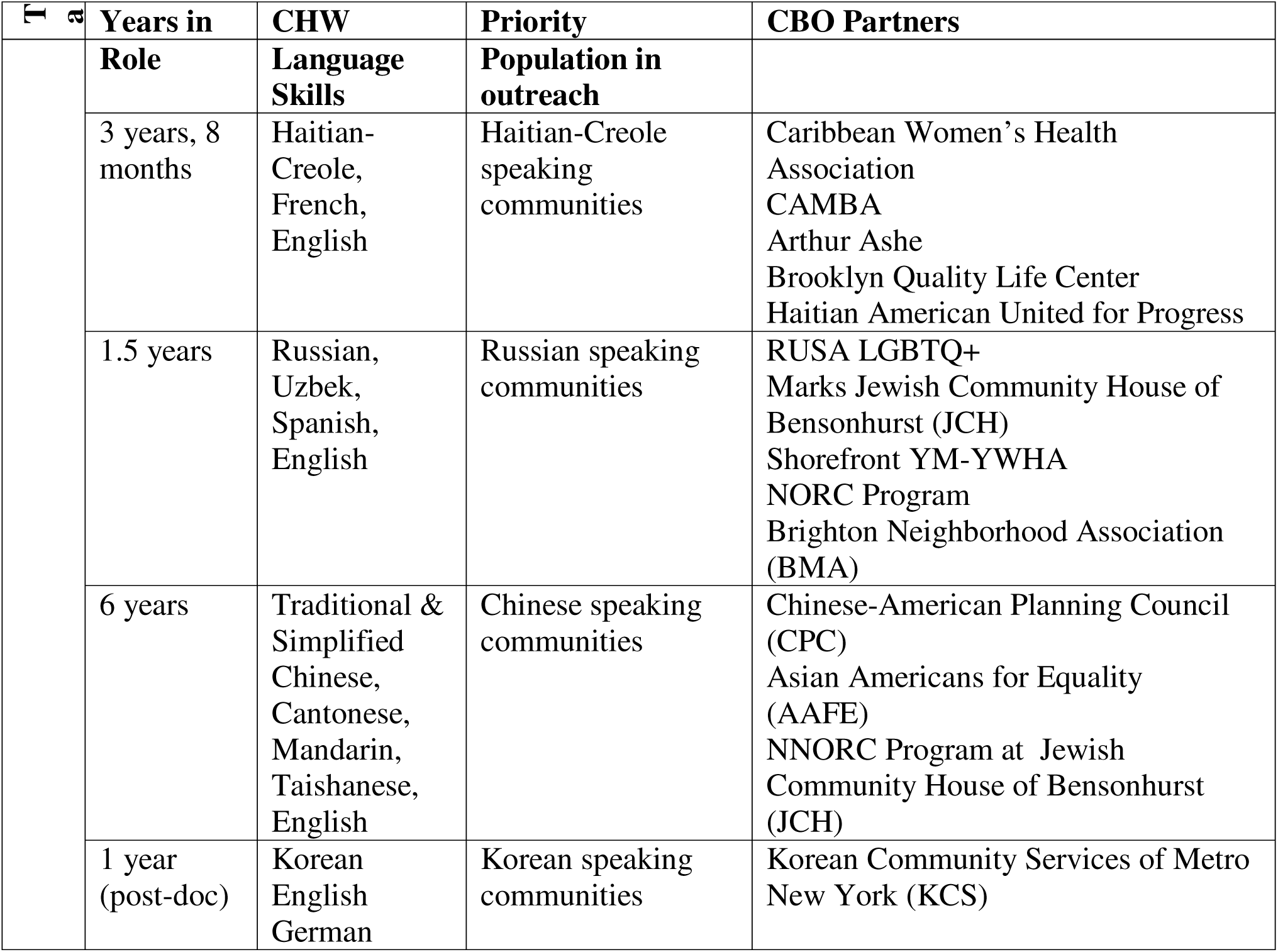
CHW Demographics.

Cancer CHRNA was designed as a large-scale, community-engaged study to identify determinants of cancer prevention and disparities, assess the role of racial discrimination in healthcare access and prevention behaviors, and document community-level health priorities among racial and ethnic minoritized and immigrant populations. A second optional module examined how structural factors affect daily life, and access to the healthcare system, and the food system.

At its foundation, Cancer CHRNA employed a community-based participatory research (CBPR) approach, engaging community partners and leveraging bilingual and bicultural CHWs to establish robust community–clinical linkages. CHWs were integral to the study team: they reviewed and refined the survey instrument, identified and affirmed CBO partners, advised on community-based recruitment sites, and led data collection. Data were collected in both clinic and community settings in nine written languages: English, Arabic, Bangla, Chinese (Simplified and Traditional), Haitian-Creole, Korean, Russian, Spanish, and Urdu (see Table 1).

### Study Methods

This study is part of a qualitative process evaluation of Cancer CHRNA and was led by an external evaluator. The evaluation aimed to understand implementation processes, particularly the role of trust, from the perspective of CHWs who led recruitment and data collection efforts.

We applied the Consolidated Framework for Implementation Research (CFIR) to guide our evaluation of implementation factors across individual, organizational, and community levels(18). As trust emerged as a dominant and cross-cutting theme, we incorporated the Theoretical model for trusting relationships and implementation(19) to further interpret the trust-building strategies used by CHWs and CBOs.

### Data Sources

Data from this analysis were drawn from three qualitative sources: 1) Field notes documented by CHWs during the implementation process, capturing real-time insights about recruitment, interactions with participants, and adaptations in outreach strategies. Field notes were used to guide in-depth interview questions. 2) In-depth interviews with four bilingual and bicultural CHWs representing the four largest linguistic communities in the study (Russian, Chinese, Haitian-Creole, and Korean). Interviews lasted 60-90 minutes and explored implementation strategies, community engagement, challenges, and reflections on trust-building. 3) A focus group with key research team members to discuss the development and implementation of Cancer CHRNA, including the structural factors module, partnerships, translation processes, and reflections on building community trust.

Interview guides and focus group protocols were semi-structured and designed to capture both planned and emergent elements of implementation. Guides are included in the Supplementary Materials.

### Participants

We purposively selected CHWs with the highest recruitment numbers and most sustained engagement in the implementation process. Table 1 provides demographic and background information on the CHWs interviewed, including their years of experience, language proficiencies, populations served, and CBO partners.

### Data Analysis

All interviews were recorded via Zoom, transcribed verbatim, and analyzed using a template analysis approach based on a priori themes of constructs from CFIR (18), while allowing for emergent themes from the data (thematic analysis).

As the theme of trust emerged repeatedly across all CFIR domains, we conducted an additional round of focused coding using Metz et al. model for trusting relationships and implementation (19). This model articulates two core mechanisms of trust-building: 1) Relational strategies (e.g., authenticity, bi-directional communication, empathy, vulnerability) aimed at strengthening the mutuality and quality of relationships; 2) Technical strategies (e.g. demonstration of expertise, frequent interactions, responsiveness) focused on demonstrating reliability, competence, and credibility (see Figure 1). This model’s theory of change suggests that the consistent use of relational and technical strategies fosters positive affect, mutual understanding, and sustained engagement, key precursors to effective implementation.

**Figure 1.**
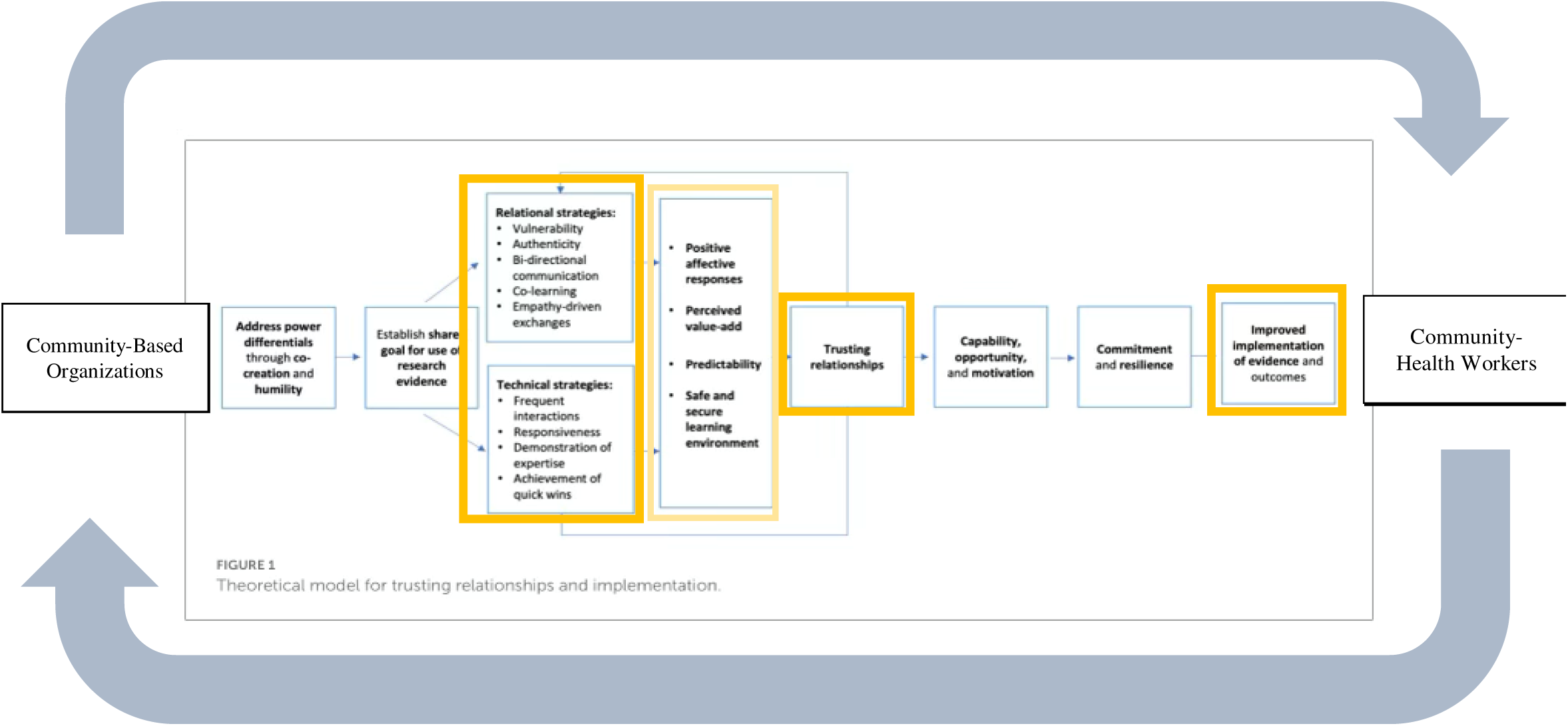
Adapted Theoretical model for trusting relationships and implementation (19)

Three trained researchers (AR, YY, and DM) independently coded each transcript using the developed codebook. We coded using the codebook, compared our code applications and assessed interrater reliability through a shared understanding and consensus on the applications and revised our codebook. Dedoose software was used for the qualitative coding process.

## Results

We analyzed four in-depth interviews with CHWs serving Russian, Chinese, Haitian-Creole, and Korean-speaking populations, as well as one focus group with the research team. Although the field notes, interviews, and focus group explored a broad range of implementation topics, trust emerged as a central theme across all linguistic and cultural groups.

Three key themes captured how trust was established and sustained during the needs assessment: 1) CHWs as trusted messengers embodying trustworthiness; 2) CBO partnerships as bridges to institutional trust; and 3) Sustaining long-term trust among CHWs, CBOs, and participants. Each theme was supported by specific relational and technical strategies aligned with Metz et al.’s theoretical model(19).

## CHWs demonstrating and embodying trustworthy attributes

CHWs served as the primary implemented of Cancer CHRNA and were central to establishing interpersonal trust with participants. Consistent with Metz et al.’s model, CHWs employed both relational strategies including authenticity, empathy, bi-directional communication, and vulnerability, and technical strategies including demonstration of expertise, frequent interactions, and responsiveness, to build and maintain trust (See Table 2). Table 2 maps each subtheme to Metz et al.’s relational and technical trust-building strategies and provides exemplar quotes from the interviews with CHWs.

**Table 2.**
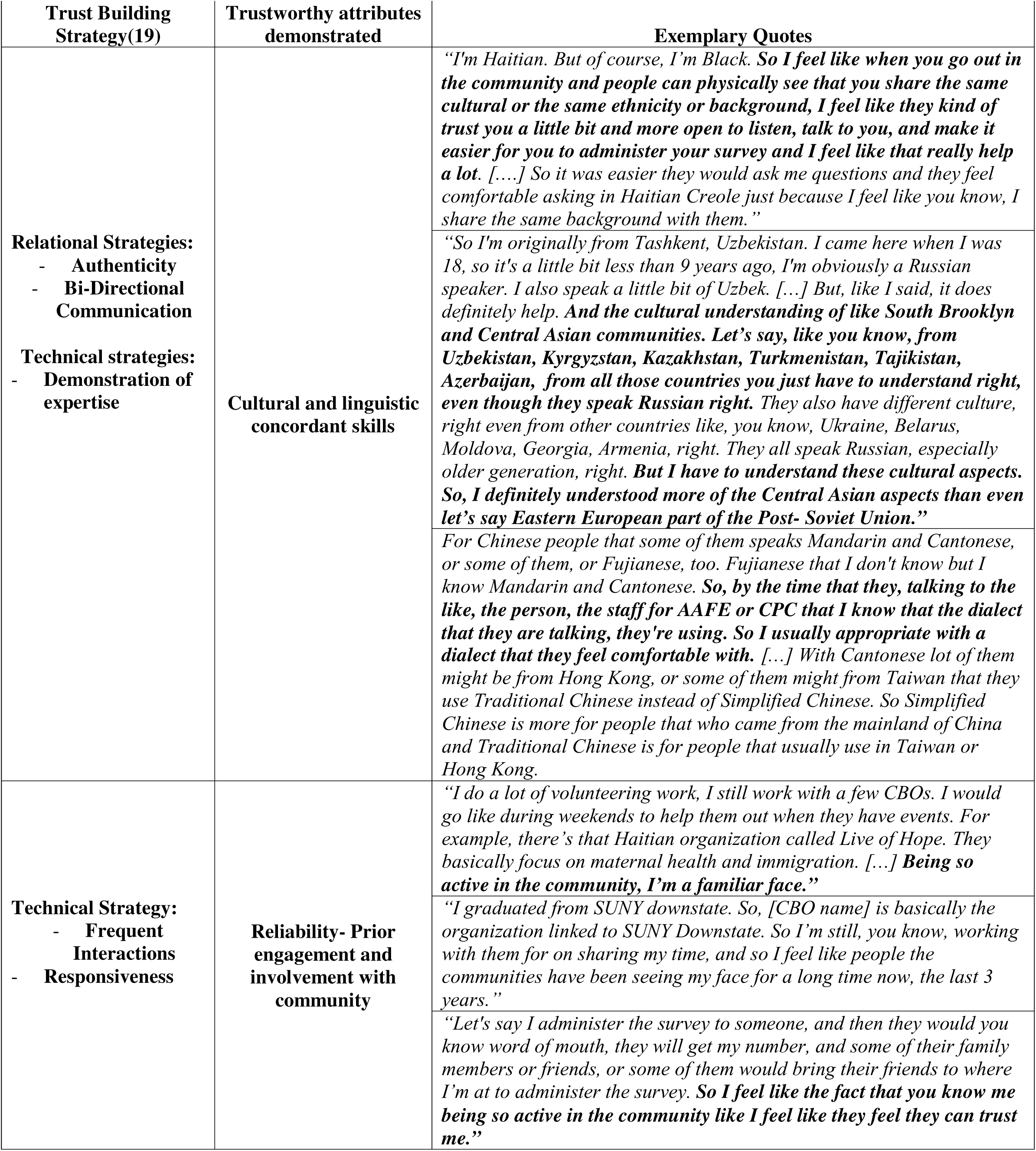
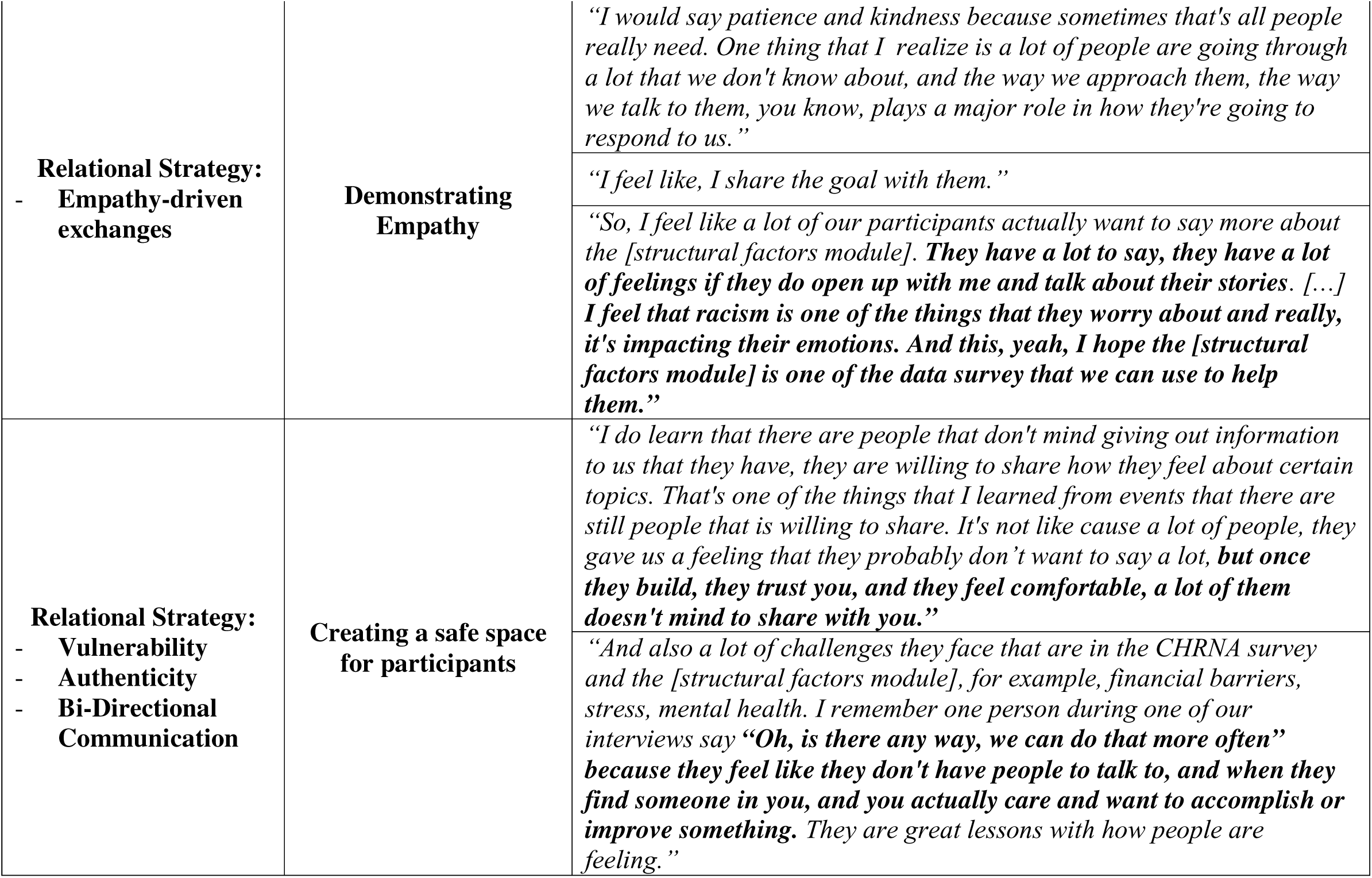
CHWs as trust messengers-demonstrating or embodying trustworthiness.

| Trust Building Strategy(19) | Trustworthy attributes demonstrated | Exemplary Quotes |
| --- | --- | --- |
| **Relational Strategies:**<br>- Authenticity<br>- Bi-Directional Communication<br>**Technical strategies:**<br>- Demonstration of expertise | **Cultural and linguistic concordant skills** | *“I’m Haitian. But of course, I’m Black. So I feel like when you go out in the community and people can physically see that you share the same cultural or the same ethnicity or background, I feel like they kind of trust you a little bit and more open to listen, talk to you, and make it easier for you to administer your survey and I feel like that really help a lot. [...] So it was easier they would ask me questions and they feel comfortable asking in Haitian Creole just because I feel like you know, I share the same background with them.”*<br>*“So I’m originally from Tashkent, Uzbekistan. I came here when I was 18, so it’s a little bit less than 9 years ago, I’m obviously a Russian speaker. I also speak a little bit of Uzbek. [...] But, like I said, it does definitely help. And the cultural understanding of like South Brooklyn and Central Asian communities. Let’s say, like you know, from Uzbekistan, Kyrgyzstan, Kazakhstan, Turkmenistan, Tajikistan, Azerbaijan, from all those countries you just have to understand right, even though they speak Russian right. They also have different culture, right even from other countries like, you know, Ukraine, Belarus, Moldova, Georgia, Armenia, right. They all speak Russian, especially older generation, right. But I have to understand these cultural aspects. So, I definitely understood more of the Central Asian aspects than even let’s say Eastern European part of the Post- Soviet Union.”*<br>*For Chinese people that some of them speaks Mandarin and Cantonese, or some of them, or Fujianese, too. Fujianese that I don’t know but I know Mandarin and Cantonese. So, by the time that they, talking to the like, the person, the staff for AAFE or CPC that I know that the dialect that they are talking, they’re using. So I usually appropriate with a dialect that they feel comfortable with. [...] With Cantonese lot of them might be from Hong Kong, or some of them might from Taiwan that they use Traditional Chinese instead of Simplified Chinese. So Simplified Chinese is more for people that who came from the mainland of China and Traditional Chinese is for people that usually use in Taiwan or Hong Kong.* |
| **Technical Strategy:**<br>- Frequent Interactions<br>- Responsiveness | **Reliability- Prior engagement and involvement with community** | *“I do a lot of volunteering work, I still work with a few CBOs. I would go like during weekends to help them out when they have events. For example, there’s that Haitian organization called Live of Hope. They basically focus on maternal health and immigration. [...] Being so active in the community, I’m a familiar face.”*<br>*“I graduated from SUNY downstate. So, [CBO name] is basically the organization linked to SUNY Downstate. So I’m still, you know, working with them for on sharing my time, and so I feel like people the communities have been seeing my face for a long time now, the last 3 years.”*<br>*“Let’s say I administer the survey to someone, and then they would you know word of mouth, they will get my number, and some of their family members or friends, or some of them would bring their friends to where I’m at to administer the survey. So I feel like the fact that you know me being so active in the community like I feel like they feel they can trust me.”* |
| **Relational Strategy:**<br>- Empathy-driven exchanges | **Demonstrating Empathy** | *"I would say patience and kindness because sometimes that's all people really need. One thing that I realize is a lot of people are going through a lot that we don't know about, and the way we approach them, the way we talk to them, you know, plays a major role in how they're going to respond to us."* |
|  |  | *"I feel like, I share the goal with them."* |
|  |  | *"So, I feel like a lot of our participants actually want to say more about the [structural factors module]. **They have a lot to say, they have a lot of feelings if they do open up with me and talk about their stories.** [...] **I feel that racism is one of the things that they worry about and really, it's impacting their emotions. And this, yeah, I hope the [structural factors module] is one of the data survey that we can use to help them.**"* |
| **Relational Strategy:**<br>- Vulnerability<br>- Authenticity<br>- Bi-Directional Communication | **Creating a safe space for participants** | *"I do learn that there are people that don't mind giving out information to us that they have, they are willing to share how they feel about certain topics. That's one of the things that I learned from events that there are still people that is willing to share. It's not like cause a lot of people, they gave us a feeling that they probably don't want to say a lot, **but once they build, they trust you, and they feel comfortable, a lot of them doesn't mind to share with you.**"* |
|  |  | *"And also a lot of challenges they face that are in the CHRNA survey and the [structural factors module], for example, financial barriers, stress, mental health. I remember one person during one of our interviews say **"Oh, is there any way, we can do that more often"** because they feel like they don't have people to talk to, and when they find someone in you, and you actually care and want to accomplish or improve something. They are great lessons with how people are feeling."* |

A foundation mechanism was cultural and linguistic concordance, which enabled CHWs demonstrated authenticity and bi-directional communication (relational strategies) while also showcasing expertise (technical strategy). As one CHW explained, speaking participants’ preferred language made it easier for them to ask questions openly and feel comfort engaging in the needs assessment.

> *“One thing that I would always get from the Haitian population now, you know many don’t trust the health care system or the government. So it was easier [when] they would ask me question[s] and feel comfortable asking in Haitian Creole just because I share the same background with them”.*

This was echoed by CHWs across communities, who described in their field notes how shared cultural knowledge helped them adapt their recruitment approaches and address deep-rooted mistrust of healthcare systems.

> *“As trust being a main issue here in such a reserved community, I developed a strategy with patients by asking them “How are you feeling today?” and introducing myself “My name is [..], I work here and I am here to make sure we can improve your healthcare experience and your benefits and eliminate any health disparities you may face” […] and offered them to proceed to a more private room so I can ask them couple questions about health topics. This strategy has given positive results.”*

Trust was furthered through reliability, expressed as frequent interactions and responsiveness with participants. Many CHWs had longstanding involvement with local CBOs or prior experience working on related health initiatives, which allowed them to maintain a visible and dependable presence in the community. These ongoing interactions signaled accountability and sustained investment, reinforcing participants’ perception of CHWs as reliable and trustworthy. Rather than relying solely on prior relationships, CHWs demonstrated reliability through their ability to follow through, adapt, and remain present throughout the implementation process.

Relational strategies such as empathy-driven exchanges also played a central role. CHWs emphasized that approaching participants with patience and kindness created a supportive environment for discussing sensitive topics included in the needs assessment (e.g. cancer screening and disparities, and structural factors). This empathic stance made participants feel understood and valued, encouraging them to share their perspectives.

Lastly, CHWs described the importance of creating a safe space for participants. By combining empathy, authenticity, vulnerability, and bi-directional communication, CHWs lowered perceived risks and fosters environments where participants felt comfortable disclosing personal experiences. Several noted that once trust was established, participants were often eager to talk more about their personal stories and experiences.

Together, these relational and technical strategies enabled CHWs to embody trustworthiness and strengthen connections with participants. In doing so, they established a baseline of interpersonal trust; partnerships with community-based organizations then amplified that trust at the organizational and community level.

## CBO Partnerships: Building and Enhancing Trust

One of the most prominent themes that emerged from the data was the importance of building strong partnerships with CBOs and how that was key to building, enhancing, and maintaining trust among participants. CBOs were longstanding and recognizable presences within their communities, and their involvement functioned as a form of organizational endorsement that reassured participants of the CHW’s and needs assessments’ credibility.

CHWs emphasized that CBOs acted as trusted sources. Their embeddedness and continued presence in the community made participants more comfortable engaging with a research initiative affiliated with them. This dynamic reflects technical strategies of frequent interactions and responsiveness, as CBOs facilitated ongoing connections between the CHWs and community members.

> *“And also I would say the CBOs some of them been in that community for so long, and have been very helpful. So, me being there, or I feel like they all work through the community and then bringing someone I feel like they kind of like, you know trust that.”*

CBOs also enhanced perceptions of reliability. Participants’ trust in CBOs transferred to CHWs, who were often introduced through these partnerships. CHWs described how participants felt more confident sharing personal information when it is clear the work was linked to a known and trusted community organization.

> *“So they should see me as a reliable person, because they know me, and they know they trust the [CPC] organization. And I am not from somewhere that it might be like a fraud or somewhere, you know, that might steal their information. This is one of the big things for Chinese immigrants. They often receive calls from nowhere, and asking about their personal information, and they saw the news that there are people that got their identities stole because of giving out information.”*

Together, CBO partnerships served as bridges between the academic health system and the community, reinforcing the credibility of CHWs and creating access points for community members who might not otherwise be engaged in healthcare. (See Table 3 for exemplar quotes).

**Table 3.** CBO partnerships to build and enhance trust within communities and among participants.

|  |  |  |
| --- | --- | --- |
| Trust Building Strategies(19) | Subthemes | Exemplary quotes |
| Technical Strategies: | CBOs as trusted | *"And also I would say the CBOs some of them been in that community for so long, and have been very helpful. So, me being* |
| - **Frequent interactions** | **sources** | *there, or I feel like they all work through the community and then bringing someone I feel like they kind of like, you know trust that.”* |
|  |  | *“It was important for us to have these relationships with community organizations, because they bring a set of individuals that they serve which is different in some ways to individuals that access health care institutions already. So having the voices of these community members that go to the trust community-based organizations but may not actually end up going to the healthcare institutions was important for us to have.” [focus group discussion]* |
| **Technical Strategies:**<br>- **Frequent interactions**<br>- **Responsiveness** | **CBO reliability** | *“And also I would say the CBOs some of them been in that community for so long, and have been very helpful. So, me being there, or I feel like they all work through the community and then bringing someone I feel like they kind of like, you know trust that.”* |
|  |  | *“So they should see me as a reliable person, because they know me, and they know they trust the [CPC] organization. And I am not from somewhere that it might be like a fraud or somewhere, you know, that might steal their information. This is one of the big things for Chinese immigrants. They often receive calls from nowhere, and asking about their personal information, and they saw the news that there are people that got their identities stole because of giving out information.”* |

## Sustaining Trust Over Time

Beyond establishing initial connections, CHWs and CBOs demonstrated how trust can be maintained and deepened across the timeline of the Cancer CHRNA. Sustained trust was evidenced in two primary ways: long-term collaboration with CBOs and continued engagement with participants.

First, CHWs described how long-term collaboration with CBOs provided stability and continuity. Many partnerships were built on years of prior work together, allowing both parties to rely on established rapport. This longevity reflected technical strategies of frequent interactions and reliability, with CHWs and CBOs maintaining contact through collaboration. These trusted relationships created pathways for recruitment and reinforced mutually beneficial relationships.

Second, CHWs emphasized how trust carried over into continued engagement with participants. Even when new module, such as the structural factors that influence healthcare questions, were introduced later in the assessment, participants who had built rapport with CHWs were willing to return and contribute additional data. This continuation was enabled by relational strategies of authenticity and technical strategies of frequent interactions.

Overall, the sustainability of trust demonstrates that relational and technical strategies were not one-time efforts but cumulative processes. By nurturing trust over time, CHWs and CBOs created conditions for ongoing collaboration between communities and institutions, with implications for future initiatives beyond Cancer CHRNA. (See Table 4 for exemplar quotes.)

**Table 4.** Sustained Trust.

| **Trust Building Strategies** | **Sub-themes** | **Exemplary quotes** |
| --- | --- | --- |
| **Relational strategies:**<br>- Authenticity<br>**Technical Strategies:**<br>- Frequent interactions | **Long-term collaboration with CBOs (sustainable trusted partnership)** | *“And these kinds of programs working together, we built up to trust and the partnerships we have partnerships with them for years.”*<br>*“I would say that partnership to those organizations that I mentioned. It just come in naturally already, because we know each other, and we've been working with each other for years. And yeah, it just feels like we are not only just like partnerships, we are friends, kind of when we go in there, and they, we feel like that we are being welcome. And yeah, and they don't, they don't hold back to promoting our program.”*<br>*“I feel like one thing that helped the fact that I'm already working with the same organizations in different projects. We kind of always keep in touch whenever they have something. Even I'm not working in a project, I always go to the events support them whenever we can bring, resources to them whenever they ask. I feel like that's how that's how it helped. So whenever in that, in our turn whenever we need something, they always there to help and support.”* |
|  | **Continued trust with CHWs and participants** | *“So on, yeah, they were pretty open, they knew me already. So, they sort of trust me, the trust is really big thing right. It has to establish this trust. You know the participant trust you, the community member trusts you, then that's the most important thing, right. So in any of those practices I just mentioned, the trust would be also a big thing.”* |

## Discussion

To our knowledge, the Cancer CHRNA is the only community-based needs assessment conducted in nine languages with community input to examine cancer-related behavioral and social priorities, offering a unique lens on the role of trust within such an initiative. Our findings underscore the centrality of trust as a mechanism that enabled meaningful participation across immigrant communities, particularly, when addressing sensitive topics such as cancer screening disparities and structural factors that influence access to health and food systems. Trust was not incidental to implementation but the process through which recruitment, engagement, and sustained collaboration became possible.

Three overarching themes illustrated how trust was built and sustained: 1) CHWs as trusted messengers embodying trustworthiness; 2) the role of CBO partnerships in building and enhancing trust with participants; and 3) sustaining trust over time. Across these themes, CHWs and CBOs employed relational strategies (authenticity, empathy, bi-directional communication, vulnerability) and technical strategies (demonstration of expertise, frequent interactions, responsiveness, reliability), consistent with Metz et al.’s theoretical model (19). These strategies activated trust, which in turn shaped participant willingness to engage and laid a foundation for continued collaboration.

Our results are consistent with prior literature demonstrating the value of cultural and linguistic concordance, long-standing community partnerships, and CHWs’ role in bridging communities and institutions (8, 11–13, 20, 21). Additionally, this study contributes several new insights to the literature. First, it shows how cultural and linguistic concordance operated simultaneously as a relational and technical strategy. Relationally, shared language and cultural background displayed authenticity and fostered bi-directional communication, making participants more comfortable sharing their perspectives. Technically, it demonstrated expertise as CHWs could accurately explain study content, address questions, and adapt their approaches to cultural norms. This dual function was particularly impactful among the diverse immigrant communities included in Cancer CHRNA where there is documented mistrust research institutions and healthcare systems, and language barriers often deter research participation (22–26). Notably, this needs assessment was the first to implement across nine different languages and incorporate cultural and linguistic concordance for all.

Second, our findings illustrate how CBO partnerships act as organizational bridges, transferring credibility to CHWs and creating access points for community members who might not otherwise be engaged with healthcare systems. Participants’ trust in CHWs was reinforced by the longstanding presence of CBOs, underscoring an interconnected trust dynamic. Third, the study highlights the sustainability of trust, with participants re-engaging in new modules and CHWs describing partnerships with CBOs as mutually beneficial. Together, these findings affirm and add nuance to Metz et al.’s model by demonstrating how relational and technical strategies can be applied in multilingual, diverse, immigrant communities.

Empathy emerged as a critical relational strategy. By approaching participants with patience and kindness, CHWs signaled respect and created a sense of safety that encouraged disclosure on sensitive topics such as cancer screening, disparities, and structural challenges experienced in accessing quality and responsive care and resources. Empathy also functioned as a technical strategy as it created an environment to express responses to challenges questions, strengthening both engagement and data quality. This finding resonates with Relational Cultural Theory, which conceptualized empathy as mutual and growth-fostering (19, 27). In practice, CHWs’ empathic approaches created reciprocal exchanges in which participants felt heard and respected, CHWs deepened their understanding of community perspectives. This reinforced trust and sustained participants’ willingness to engage. This empathetic approach was particularly important when working with immigrant communities, many of whom historically have higher levels of mistrust towards healthcare systems and institutional research programs (22–26).

The significance of long-standing partnerships with CBOs aligns with CBPR findings which emphasize the value of existing community networks (28, 29). CBOs consistently provide community members continuity of trust through sustained engagement, communication, services, and advocacy (8, 30). The utilization of robust community networks was a key factor in the successful recruitment in Cancer CHRNA. Reliability prominently emerged in relation to both CHWs and CBOs. CHWs were perceived as reliable due to their established presence within the community. Moreover, participants’ perception of CHWs’ reliability was further enhanced by the CHWs’ affiliations with trustworthy CBOs. This interconnected trust dynamic suggests that the reliability of CBOs, recognized through their consistent community engagement and advocacy, was instrumental in augmenting the perceived reliability of CHWs. These results are consistent with CBPR literature and studies discussing increased credibility and sustainability when partnering with CBOs to address disparities (8, 29, 31–33).

The results highlight several implications for the design and implementation of future community-based needs assessments and interventions. Trust must be recognized as a core mechanism of implementation, a culmination of intentional use of relational and technical strategies, rather than a background condition (19). Central to this process is the role of CHWs, who should be positioned as key implementers of trust-building by leveraging cultural and linguistic concordance to foster authentic connections with participants. At the same time, partnerships with CBOs are essential, as these organizations act as bridges between communities and institutions, extending credibility to CHWs and facilitating access to populations who might otherwise remain underserved. Finally, sustaining trust requires long-term commitment: trust is cumulative, built through mutually beneficial collaboration, frequent interactions, and ongoing engagement that extends well beyond a single project.

### Future Directions

Maintaining trust beyond the needs assessment requires active dissemination (29). Following Cancer CHRNA, study reports were shared with participating communities, CBOs, and community advisory boards. Continued transparency and reporting back of findings are essential for sustaining trust and adhering to CBPR principles (28, 29). Future research should build on these practices, embedding relational and technical strategies into the design of interventions that aim to build and sustain trust, reduce cancer disparities, and advance health equity among immigrant populations.

### Strengths and Limitations

Our study had many strengths. First, to our knowledge, Cancer CHRNA is the first large-scale community-based cancer screening needs assessment focused on multiple priority groups and translated into multiple languages. This is particularly relevant given the disparities in cancer screening among immigrant populations in NYC (14, 15, 34). This study provides specific insights into successful trust building relationships among CBOs and participants across various diverse communities. Our study does include some limitations. First, we only interviewed four of the eight CHWs involved in implementation process as these were the full time CHWs and communities with the highest recruitment for the survey. The four CHWs included had greater collaborative involvement in each stage of the needs assessment but especially, in the implementation process. As such we were able to reach thematic saturation with the interviews conducted. Second, we did not interview Cancer CHRNA participants or CBO partners directly, but rather participant and CBO perceptions and feedback were reported by CHWs. However, our goal was to understand the perspectives of those who were engaged in the implementation process and their perspectives of the trust building process. These findings would be strengthened by the inclusion of additional diverse perspectives, learning about trust directly from community members as well as CBO partners.

## Conclusion

This study demonstrates that the success of Cancer CHRNA was rooted in the ability of CHWs and CBOs to build and sustain trust among diverse communities. Through relational strategies such as empathy, authenticity, bi-directional communication, and technical strategies such as frequent interactions, responsiveness, and demonstration of expertise, CHWs embodies trustworthiness and created safe spaces for participation. Partnerships with longstanding CBOs further enhanced credibility and opened access to community members who might not otherwise engage in academic or healthcare systems. Sustained, mutually beneficial relationships with both CBOs and participants ensured continued engagement throughout the assessment.

By highlighting how trust operated as the mechanism that made recruitment and participation excel, this study contributes practical insights for the design of future community-based needs assessments. Centering CHWs and CBOs in recruitment is not only effective but essential for building trust in diverse, multilingual communities and ensuring meaningful community engagement in research.

## Declarations

**Ethics approval and consent to participate**

**Consent for publication Not applicable.**

**Availability of data and materials**

Data are available upon request.

## Authors’ Contributions

Conceptualization of Cancer CHRNA was led by Simon Kwon in collaboration with the PCC Cancer Action Network (CAN). Implementation of Cancer CHRNA was led by CHWs, overseen by Victoria Foster. Abiha Kazmi and Yousra Yusuf were involved in data collection of Cancer CHRNA. Sonia Sifuentes managed program tracking of Cancer CHRNA. Ashlin Rakhra conducted the qualitative interviews of CHWs and focus group among the research team reported in this manuscript, performed data analysis, and wrote the manuscript. Yousra Yusuf and Deborah Min also performed data analysis. No authors had access to information that could identify individual participants during or after data collection. Simona Kwon and Victoria Foster edited the paper. All authors approved the final manuscript.

## Data Availability

All data produced in the present study are available upon reasonable request to the authors.

## ACKNOWLEDGEMENTS

The Research reported in this was supported in part by the National Cancer Institute of the National Institutes of Health award number P30CA016087, the Centers for Disease Control and Prevention award number U01DP006643, and the Dune Road Foundation, Inc.

The authors would also like to thank the following Community Health Workers, Support Staff, Interns, and Community-Based Organizations for their contribution and commitment to Cancer CHRNA and this research.

*CHWs*: Alzaharaa Ahmed, Alice Liang, Alex Trifonov, Cecelia Chabani

*Interns and support staff:* Abiha Kazmi, Deborah Min, Kimberly Charlies, Shumi Ahkter, Sumbal Marry, Yaena Song, Yousra Yusuf

*PCC Cancer Action Network (CAN):*

AMERICAN CANCER SOCIETY

AMERICAN ITALIAN CANCER FOUNDATION

ARAB AMERICAN ASSOCIATION OF NY

ARAB AMERICAN FAMILY SUPPORT CENTER

ARTHUR ASHE INSTITUTE FOR URBAN HEALTH

ASIAN AMERICANS FOR EQUALITY

BEDFORD STUYVESANT RESTORATION CORPORATION: RESTORATION PLAZA

BETTYS BREAST CANCER FOUNDATION

BLACK HEALTH MATTERS

BROOKLYN CENTER FOR QUALITY LIFE

BROOKLYN CHINESE-AMERICAN ASSOCIATION

BROOKLYN COMMUNITY PRIDE CENTER

BROOKLYN PUBLIC LIBRARY

CALLEN-LORDE

CAMBA

CANCER NETWORK

CARIBBEAN EQUALITY PROJECT

CENTER FOR FAMILY LIFE

CHEEKY CHARITY

COJECO

EDITH AND CARL MARKS JEWISH COMMUNITY HOURS OF BENSONHURST

ERASE RACISM NY

FAMILY & CHILDREN’S ASSOCIATION

FAMILY SERVICE LEAGUE, LI

FIFTH AVE COMMITTEE

FOOD BANK FOR NYC

FORT GREENE COUNCIL

GRAND STREET SETTLEMENT

HEALTHFIRST

HISPANIC COUNSELING CENTER

HOMECREST

HOPE FOR PINK

ISLAND HARVEST

JASA

KOREAN COMMUNITY SERVICES

LA JORNADA

LGBT NETWORK

MAKE THE ROAD NY

MEXICAN COALITION

MIXTECA

MORIAH CITY CHURCH

NASSAU COUNTY DEPARTMENT OF HEALTH

NASSAU COUNTY POLICE

NATIONAL SOCIETY OF HEALTH COACHES

NEW IMMIGRANT COMMUNITY EMPOWERMENT

THE NEW YORK COMMUNITY TRUST

NYC DEPARTMENT FOR THE AGING

NYC DEPARTMENT OF HEALTH AND MENTAL HYGIENE

OFFICE OF THE NYC PUBLIC ADVOCATE

ONE LIFE CHRISTIAN CHURCH

OUTREACH NEW YORK

PLANNED PARENTHOOD OF GREATER NY

PUBLIC HEALTH SOLUTIONS

QARAVAN

RAINBOW HEIGHTS

RAISINGHEALTH

THE RETREAT

RISEBORO

ROCKVILLE CENTRE BREAST CANCER COALITION

SALAM ARABIC CHURCH

SALVATION ARMY

SEPA MUJER

SHARE

SHORE FRONT YM-YWHA

SUFFOLK COUNTY DEPT. OF HEALTH SERVICES

TRANSLATINX NETWORK

UNIVERSITY SETTLEMENT

WOMEN’S EMPOWERMENT COALITION OF NY

YEMENI AMERICAN MERCHANTS ASSOCIATION

YES COMMUNITY COUNSELING CENTER

